# Predicting COVID-19 pandemic waves with biologically and behaviorally informed universal differential equations

**DOI:** 10.1101/2023.03.11.23287141

**Authors:** Bruce Kuwahara, Chris T. Bauch

## Abstract

In the early stages of the COVID-19 pandemic, it became clear that pandemic waves and population responses were locked in a mutual feedback loop. The initial lull following strict interventions in the first wave often led to a second wave, as restrictions were relaxed. We test the ability of new hybrid machine learning techniques, namely universal differential equations (UDEs) with learning biases, to make predictions in a such a dynamic behavior-disease setting. We develop a UDE model for COVID-19 and test it both with and without learning biases describing simple assumptions about disease transmission and population response. Our results show that UDEs, particularly when supplied with learning biases, are capable of learning coupled behavior-disease dynamics and predicting second waves in a variety of populations. The model predicts a second wave of infections 55% of the time across all populations, having been trained only on the first wave. The predicted second wave is larger than the first. Without learning biases, model predictions are hampered: the unbiased model predicts a second wave only 25% of the time, typically smaller than the first. The biased model consistently predicts the expected increase in the transmission rate with rising mobility, whereas the unbiased model predicts a decrease in mobility as often as a continued increase. The biased model also achieves better accuracy on its training data thanks to fewer and less severely divergent trajectories. These results indicate that biologically informed machine learning can generate qualitatively correct mid to long-term predictions of COVID-19 pandemic waves.

**Significance statement:** Universal differential equations are a relatively new modelling technique where neural networks use data to learn unknown components of a dynamical system. We demonstrate for the first time that this technique is able to extract valuable information from data on a coupled behaviour-disease system. Our model was able to learn the interplay between COVID-19 infections and time spent travelling to retail and recreation locations in order to predict a second wave of cases, having been trained only on the first wave. We also demonstrate that adding additional terms to the universal differential equation’s loss function that penalize implausible solutions improves training time and leads to improved predictions.

## Introduction

The COVID-19 pandemic generated an enormous demand for mathematical models to predict cases and guide policy [1, 2, 3]. These models were often mechanistic in nature, seeking to represent known or hypothesized transmission mechanisms in a mathematical, stochastic, or agent-based framework [4, 5, 6]. The advance warning provided by these models allowed public health institutions to prepare by implementing policies to mitigate the second wave when it arrived [7, 8]. Modelling efforts were widely applied to investigate the impact of public health measures such as testing [9], school and workplace closures [10, 11], vaccination strategies [12, 13, 14] or to stimulate action by projecting the impacts of a worst case ‘do nothing’ scenario where governments and populations did not attempt to mitigate the pandemic [8, 10].

Fortunately, most governments and members of the public did respond to the pandemic, by taking measures to reduce case incidence. Numerous studies have shown that non-pharmaceutical interventions such as lockdowns, school closures, and social distancing protocols reduce case notifications and health impacts of COVID-19 [15, 16, 17, 18]. The anticipation of infection risk in the face of rising case incidence supports adherence to these measures [19]. However, as risk of infection wanes, so too does willingness to abide by these mentally fatiguing [20, 21] and economically costly measures [21]. The relaxation of mitigation efforts may then potentially result in another pandemic wave. This two-way interaction–where infection spread influences behaviour, which influence infection spread, in turn–suggests the concept of coupled behavior-disease systems [22] may be useful for studying COVID-19 pandemic waves.

As such, many mechanistic models informed by economic, social or psychological assumptions have incorporated behaviour-disease dynamics to study the impact of interventions in the context of population behavioural feedbacks [23, 24, 25, 26, 27, 28, 29], including for COVID-19 [30, 12, 31, 32, 33]. Among the most valuable insights provided by these models is the occurrence of multiple pandemic waves, which are predicted under a wide range of conditions by these models due to waning stringency causing a resurgence of the infection [34, 12, 35, 36]. With hindsight, we can confirm that these models were correct – second waves occurred virtually everywhere during the COVID-19 pandemic (and before the arrival of new variants).

Alongside these mechanistic models, the plethora of epidemiological, sociological, mobility, and economic data generated by the pandemic allowed machine learning models to flourish [37, 38, 39, 40, 41]. These models have proven adept at integrating vast quantities of data on a multitude of factors (including behaviour) affecting disease spread. Consequently, they often adapt better to regional variability compared to mechanstic models [41, 40]. However, machine learning models have significant drawbacks. They are able to fit existing data well and accurately predict days to a couple of weeks into the future, but pay for this predictive accuracy with reduced interpretability compared to traditional models [37]. Compared to mechanistic models with relatively few easily-understood parameters, it is far more difficult to extract qualitative understandings of disease dynamics (such as second waves) from the hundreds or thousands parameters in purely machine-learning models. They are also easy to over-fit (although mechanistic models also suffer form this risk), meaning their predictive value may be limited.

Recently, advances in high-performance automatic differentiation have enabled new techniques that combine the interpretability and qualitative understanding from mechanistic models with the potentially higher predictive power and scalability of machine learning. Physics-informed machine learning (PIML) is one such methodology. The key idea is to create ML models that encode physical laws by inferring them from large amounts of data (observational bias), building them into the model’s architecture (structural bias), or training the model to uphold them (learning bias) [42]. Of particular interest for qualitative epidemic modelling are the latter two biases, as they reduce the model’s reliance on large amounts of data. By incorporating these biases, the model is prevented (in the case of structural biases) or at least discouraged (for learning biases) from making biologically impossible predictions. Learning biases can also discourage overfitting the data by introducing other objectives for the model.

The majority of learning bias applications thus far have been solving various forms of partial differential equations (PDEs) [43, 44, 45]. In these models, a neural network is trained to simultaneously fit data and to satisfy a PDE. In addition to physics, learning biases have been used in biologically-informed machine learning (BIML) applications. These include blood flow dynamics [46], drug responses [47], and cancer detection and classification [48, 49, 50]. This approach has yet to be applied to COVID-19 modeling, but has seen great success for solving large and complex partial differential equations.

A novel technique for structural bias is universal differential equations (UDEs). This method involves training neural networks embedded in differential equation models. Known dynamics can be included explicitly, while leaving unknown processes to be learned by the neural network [51]. The explicit parts of the UDE can be made to retain valuable laws such as invariant quantities. UDEs have been applied successfully on predator-prey models, metabolic networks, batteries, and photonics [51, 52]. For instance, recent research uses a neural network to learn the change in COVID-19 quarantine measures in a population over time, within the framework of a modified QSEIR (quarantine, susceptible, infectious, recovered) model. The trained network was then used to quantify the effectiveness of those measures for different regions [39, 53].

The ability of hybrid machine learning approaches – such as UDEs and PIML – to make qualitative long-term predictions about epidemic dynamics of the sort provided by mechanistic models has not yet been widely tested. This motivated our research: our objective was to combine observational biases (UDEs) with satisfiable learning biases in a coupled behaviour-disease model for COVID-19. We trained a compartmental UDE model to fit behavioural and epidemic data while penalizing deviations from several simple socio-biological assumptions. We hypothesized that a UDE model can learn the pattern of behaviour-disease interactions and hence predict a second wave (either qualitatively or quantitatively), having only seen the first wave (and its learning biases). We also hypothesized that without those learning biases, the model will learn much less effectively.

## Results

A complicated mathematical model can easily be made to fit an epidemic curve, but runs the risk of over-fitting the data and thus not being useful for prediction [54, 55]. Simpler mathematical models allow us to test our hypotheses by incorporating aspects we understand without becoming overburdened by details that we cannot reliably describe mathematically [55].

Hence, we used a UDE framework that allows us to leave the behaviour-disease dynamics of a simple compartmental behaviour-disease model unspecified, save for a few plausible assumptions (“learning biases”). In doing so, we can test the validity of those assumptions. Compartmental models divide the human population into mutually exclusive compartments based on infectious status, and which are generally implemented as differential equations. These models have been a mainstay of mathematical epidemiology for decades [5].

The algorithm learned the manner in which force of infection responds to mobility and manner in which mobility responds to its current value, the number of active cases, recent new cases, and recovered cases. The learning biases inform the model with several plausible assumptions: namely that force of infection increases with mobility, that mobility decreases with more active and recent cases, that mobility tends toward 0 (the pre-COVID average) in the absence of cases, that this tendency is stronger the more people have recovered, and that mobility cannot fall below a 100% reduction or exceed a 200% increase from the pre-COVID average (see Methods). The learning biases strongly discourage infeasible values of mobility and make data-fitting relatively less important for the optimizer. As a result, the model makes out-of-sample predictions (i.e. second waves) frequently.

In order to ensure consistent and repeatable results, we ran the model on each region 100 times both with and without learning biases. We trained the algorithm on the first wave and tested whether it could predict the second wave. Overall, the model with learning biases was successful in every region in which we tested it, though some more so than others. It consistently learned to fit the data and constraint losses, predicted second waves, and seldom made biologically implausible predictions.

We compared UDE models with and without learning bias. The model without learning biases, while not entirely a failure, was much less successful. Though it was generally able to fit the data, it predicted second waves much less frequently and made many more unrealistic predictions. Details are provided in the following subsections.

### Model Predictions

To get a sense of the model’s average behaviour, we plotted the median prediction of the 100 simulations for each region. An example for New York can be found in Section 2, Figure 2 (analogs for other regions can be found in the Supplementary Appendix, figures 3-13). The model with learning biases has consistent behaviour within the training region. The median prediction shows a small second wave and the inter-quartile range shows one of similar size to the first. The model without learning biases fits the data comparably well, but has greatly reduced variability outside of the training region. Second wave predictions are smaller or non-existent, typically only suggested by the upper quartile rather than the median. Section 2.1 Table 1 gives a numerical summary of the model’s performance across all regions. Section 2.1 Figure 1 shows a graphical summary of model performance across all regions.

**Fig. 1.**
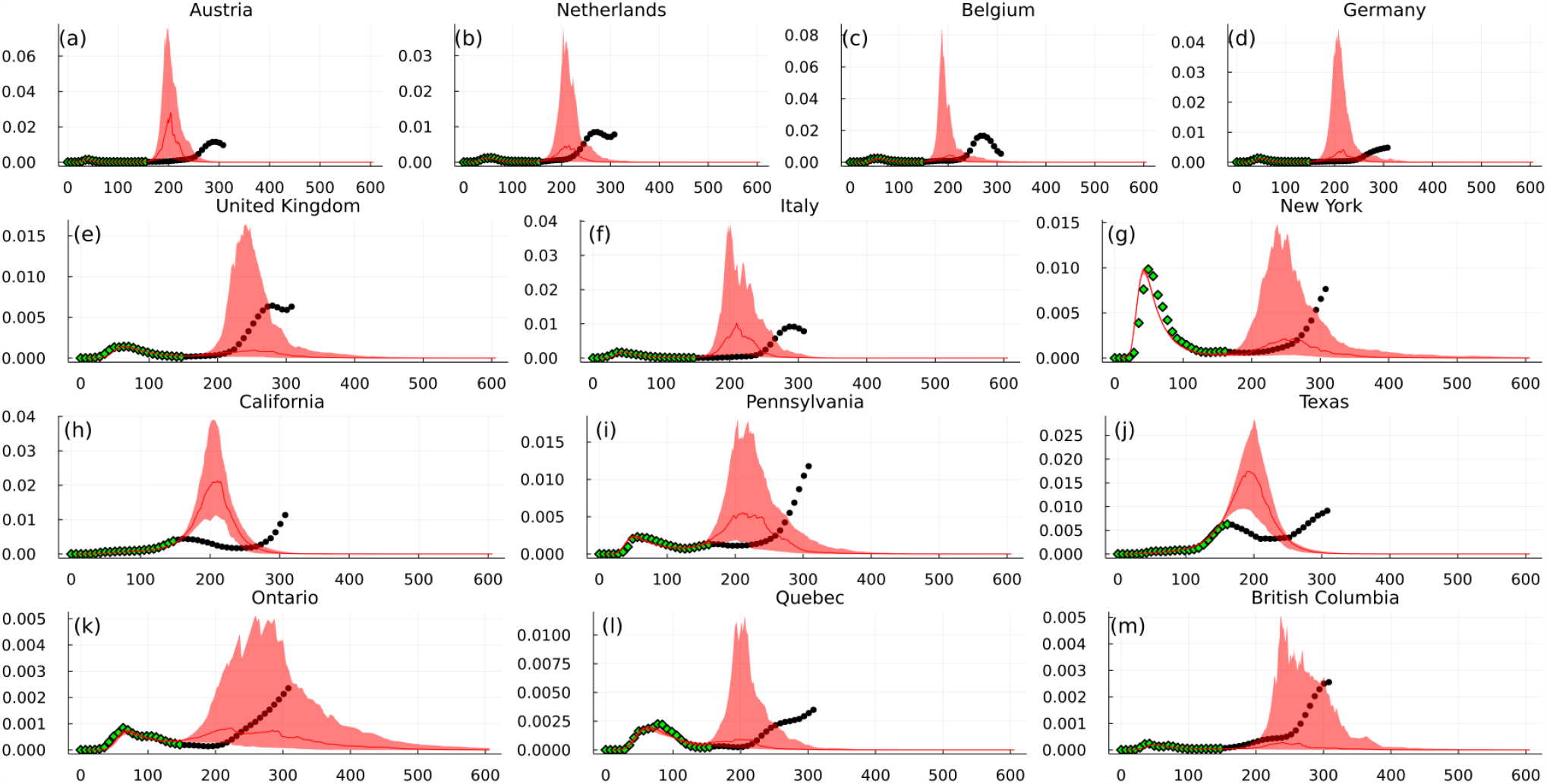
Infection prevalence time series predictions for all regions produced by the model with learning biases. Infection prevalence is the proportion of the total population that is infected at any given time. Green dots represent training data (first 22 weeks) and black dots show unseen data (a further 23 weeks). Predictions are generated using the median (solid line) and interquartile range (ribbon) of 100 independently-trained instances of the model per region.

**Fig. 2.**
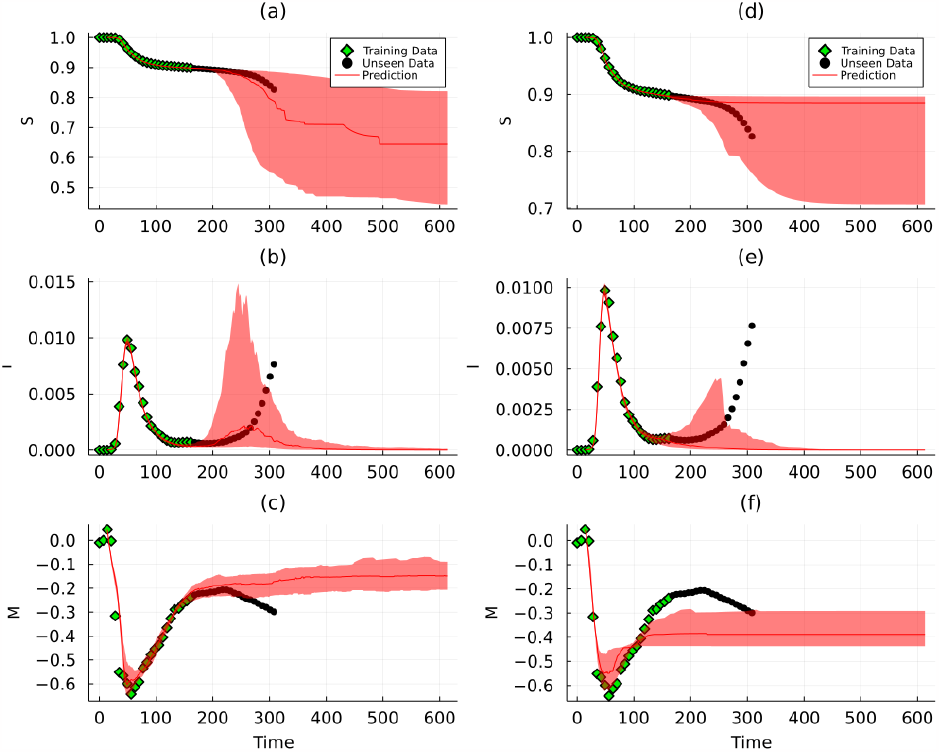
Predicted time series of all model states for New York state with learning biases (a-c) and without (d-f). Panels (a) and (b) show susceptible fraction, (b) and (d) show infected, and (c) and (f) show mobility. Green dots represent training data, while black dots represent unseen data. All predictions are generated using the median (solid line) and interquartile range (ribbon) of 100 independently-trained instances of the model.

**Fig. 3.**
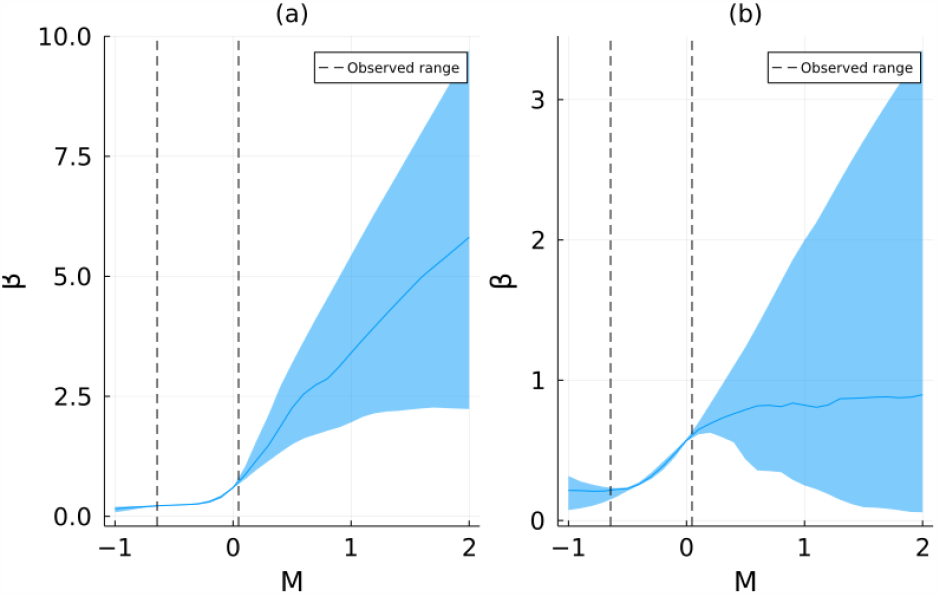
Predicted force of infection based on mobility level for New York state with learning biases (a) and without (b). Dotted lines indicate values of mobility seen by the model during training. Solid line shows the median prediction of 100 model instances and ribbon shows interquartile range.

**Table 1.** Summary of second wave predictions: Listed as biased model / unbiased model

| Region | True wave time<sup>1,2</sup> | True wave size<sup>1</sup> | 2nd wave rate | Mean wave timing<sup>1,3</sup> | Mean wave size |
| --- | --- | --- | --- | --- | --- |
| Austria | 294 | $1.15 \times 10^{-2}$ | 0.86 / 0.50 | 222 / 197 | $5.88 \times 10^{-2}$ / $2.67 \times 10^{-2}$ |
| Belgium | 273 | $1.67 \times 10^{-2}$ | 0.68 / 0.23 | 217 / 185 | $5.18 \times 10^{-2}$ / $5.71 \times 10^{-3}$ |
| Germany | 315 | $4.90 \times 10^{-3}$ | 0.78 / 0.45 | 259 / 244 | $4.53 \times 10^{-2}$ / $2.87 \times 10^{-2}$ |
| Netherlands | 273 | $8.50 \times 10^{-3}$ | 0.68 / 0.23 | 226 / 119 | $4.09 \times 10^{-2}$ / $2.43 \times 10^{-3}$ |
| Italy | 287 | $9.18 \times 10^{-3}$ | 0.83 / 0.27 | 214 / 151 | $4.46 \times 10^{-2}$ / $6.47 \times 10^{-3}$ |
| United Kingdom | 280 | $6.44 \times 10^{-3}$ | 0.63 / 0.40 | 266 / 140 | $2.95 \times 10^{-2}$ / $1.16 \times 10^{-3}$ |
| California, USA | 336 | $1.85 \times 10^{-2}$ | 0.17 / 0.010 | 454 / 228 | $2.74 \times 10^{-3}$ / $2.91 \times 10^{-4}$ |
| New York, USA | 350 | $1.26 \times 10^{-2}$ | 0.79 / 0.39 | 276 / 190 | $1.94 \times 10^{-2}$ / $6.42 \times 10^{-3}$ |
| Pennsylvania, USA | 329 | $1.29 \times 10^{-2}$ | 0.80 / 0.19 | 212 / 93 | $2.05 \times 10^{-2}$ / $1.38 \times 10^{-3}$ |
| Texas, USA | 217 | $3.27 \times 10^{-3}$ | 0.25 / 0.021 | 342 / 183 | $9.89 \times 10^{-3}$ / $2.45 \times 10^{-4}$ |
| British Columbia, Canada | 308 | $2.56 \times 10^{-3}$ | 0.047 / 0 | 357 / NA | $2.52 \times 10^{-3}$ / 0 |
| Ontario, Canada | 343 | $3.54 \times 10^{-3}$ | 0.098 / 0.29 | 158 / 300 | $4.19 \times 10^{-3}$ / $5.34 \times 10^{-4}$ |
| Quebec, Canada | 336 | $4.84 \times 10^{-3}$ | 0.48 / 0.51 | 534 / 73 | $1.79 \times 10^{-2}$ / $2.97 \times 10^{-3}$ |

<sup>1</sup>Calculated using the same mechanism as in section 4.3
<sup>2</sup>Measured in days since February 18, 2020
<sup>3</sup>Excluding simulations which do not predict second waves

### Biological Feasibility

Both models, with and without learning biases, tended to make feasible predictions, in the sense that all model states remained within their respective bounds. The biased model was stable 88% of the time, while the unbiased model was stable 85% of the time, across all regions.

However, when evaluating the learning bias loss functions on the trained models, it becomes clear that the model with learning biases is more reliable in this regard. The biased model achieves better losses across all loss objectives, including accuracy, compared to the unbiased model. Comparison of all loss functions can be found in the supplementary material.

The unbiased model does particularly poorly on the mobility upper and lower bounds (on the order of 10^4^ times worse than biased), and tendency for mobility to return to baseline in the absence of infection (roughly 10^3^ times worse).

### Second wave prediction

As a more robust metric for second wave prediction, we counted the number of local maxima exceeding at least 10^*−*3^ in the infected time series for each model simulation. With learning biases, the model predicted second waves regularly for most regions.

The unbiased model, meanwhile, rarely predicted second waves (Section 2.1, Table 1). Its best performance was on Quebec, where it predicted second waves 51% of the time, This was also the only region in which it outperformed the biased model, which predicted second waves 48% for of the time. Otherwise, it predicted second waves less than 66% as often as the biased model, sometimes as little as 1.6% as often. It predicted no second waves at all for British Columbia.

Most of the time, both models predict the second wave too early (exceptions being Texas and Quebec), but the biased model’s estimate is usually closer (only excepting Texas and Quebec). In terms of wave size, the biased model typically overestimates while the unbiased model shows no clear pattern.

### Transmissibility

One of the main uses of this model is that the trained neural network representing the force of infection can, once trained, be analyzed to examine the learned relationship between mobility and the transmission rate.

Section 2.4 Figure 3 shows the distribution in the response of *β* to mobility predicted by the model with learning biases for New York. The models all converge on the same relationship within the training region and on low out-of-sample values, but they diverge for large ones. It is also worth noting that the prediction is, as expected, monotonically increasing. Once again, all regions demonstrate similar behaviour (see Supplementary Appendix figures 14-24).

As with the time series predictions, the model without learning biases fits the data about equally well within the region on which it has been trained. However, outside that region, it extrapolates a flatter curve that is about equally likely to to be higher or lower than the median.

For a quantitative sense of how *β* responds to mobility, we evaluated each trained network at the baseline value of mobility to determine the value of *β*, and hence *R*_0_ (= *β/γ*), the basic reproduction number of the virus at the baseline value of mobility. We also applied Newton’s method to the trained neural network to find the value of mobility (*M*_*crit*_) at which *R*_0_ drops below 1, the value below which the infection will die out [56]. Results for the biased model can be found in table Section 2.4 Table 2. The unbiased model results are negligibly different for the *R*_0_, The results for *M*_*crit*_ are more variable. These unbiased model results can be found in the Supplementary Appendix Table 1.

**Table 2.**
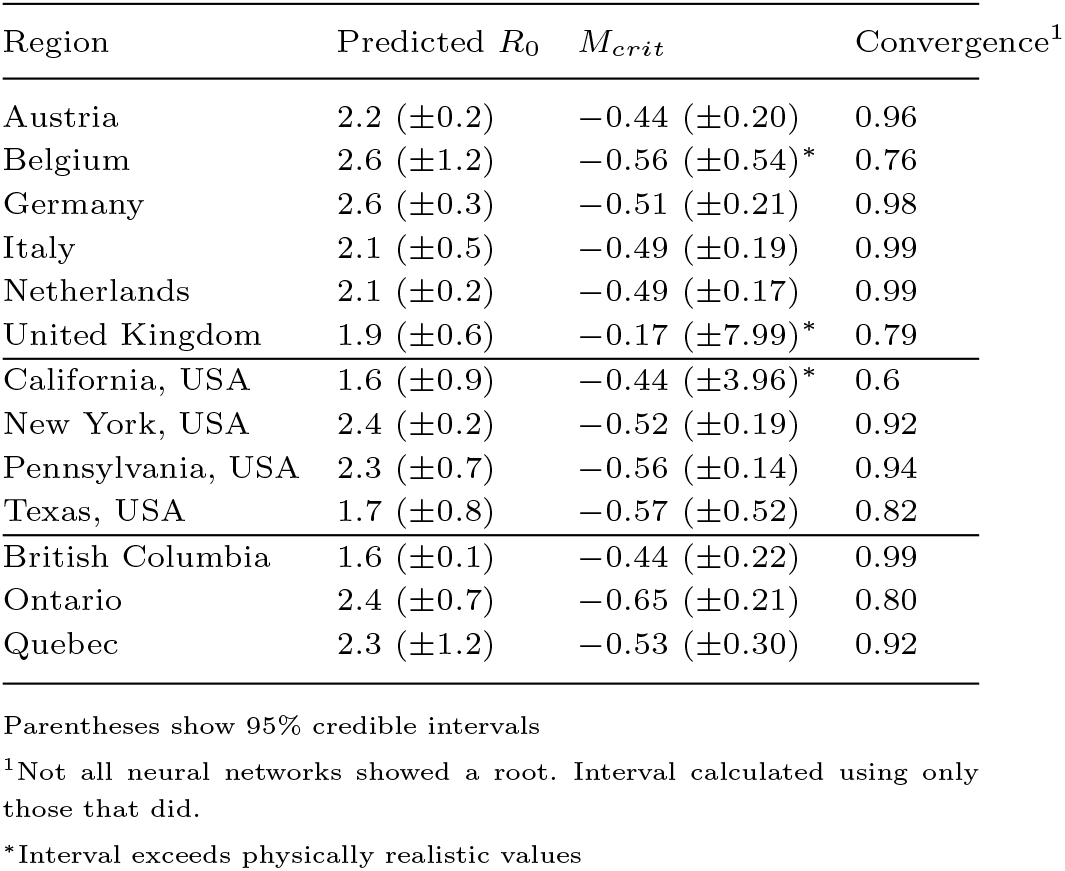
Predicted *R*_0_ and required mobility reduction

| Region | Predicted $R_0$ | $M_{crit}$ | Convergence<sup>1</sup> |
| --- | --- | --- | --- |
| Austria | 2.2 ($\pm 0.2$) | -0.44 ($\pm 0.20$) | 0.96 |
| Belgium | 2.6 ($\pm 1.2$) | -0.56 ($\pm 0.54$)* | 0.76 |
| Germany | 2.6 ($\pm 0.3$) | -0.51 ($\pm 0.21$) | 0.98 |
| Italy | 2.1 ($\pm 0.5$) | -0.49 ($\pm 0.19$) | 0.99 |
| Netherlands | 2.1 ($\pm 0.2$) | -0.49 ($\pm 0.17$) | 0.99 |
| United Kingdom | 1.9 ($\pm 0.6$) | -0.17 ($\pm 7.99$)* | 0.79 |
| California, USA | 1.6 ($\pm 0.9$) | -0.44 ($\pm 3.96$)* | 0.6 |
| New York, USA | 2.4 ($\pm 0.2$) | -0.52 ($\pm 0.19$) | 0.92 |
| Pennsylvania, USA | 2.3 ($\pm 0.7$) | -0.56 ($\pm 0.14$) | 0.94 |
| Texas, USA | 1.7 ($\pm 0.8$) | -0.57 ($\pm 0.52$) | 0.82 |
| British Columbia | 1.6 ($\pm 0.1$) | -0.44 ($\pm 0.22$) | 0.99 |
| Ontario | 2.4 ($\pm 0.7$) | -0.65 ($\pm 0.21$) | 0.80 |
| Quebec | 2.3 ($\pm 1.2$) | -0.53 ($\pm 0.30$) | 0.92 |

Parentheses show 95% credible intervals
<sup>1</sup>Not all neural networks showed a root. Interval calculated using only those that did.
\*Interval exceeds physically realistic values

The *R*_0_ predictions, averaged over all simulations for a given region, range from 1.60 (British Columbia) to 2.60 (Germany). While estimates of *R*_0_ for COVID-19 vary significantly between countries and times, this is in line with estimates of between 2.4 and 2.4 for the original COVID-19 strain [57, 58, 59, 60]. It is also consistent with other models, which found Germany and the Netherlands to have higher values [61].

The model typically estimates that a 40-50% reduction in mobility is necessary to reduce *R*_0_ below 1. This is consistently more extreme than other studies have found (20-40%) [62], but not entirely implausible considering the interquartile range. That said, we cannot interpret any result for Belgium, California, or the UK where the interquartile range exceeds physically realistic bounds.

## Discussion

Our results show that socially and biologically informed machine learning models can perform qualitative prediction tasks. When supplied with learning biases, the model routinely predicted second pandemic waves similar to those that occurred in most populations during the COVID-19 pandemic. The model seldom produced implausible predictions for mobility, and where it did, this tended to result from a failure to converge during training.

The most significant result is that the biased model predicts a second wave in every region except the Canadian province of British Columbia. The biased model predicted the second wave peak more consistently and closer to the actual time than the unbiased model without behavioural (mobility) feedback. The biased model also tended to predict a second wave that was much larger than the first wave, as occurred in most populations during the COVID-19 pandemic, although the predicted second wave was often larger in magnitude than what occurred in reality.

This ability to predict second waves is valuable from a public health perspective, for mitigation of population health impacts. Though our model does not explicitly include government policy, it can influence behaviour, and knowing the likely trajectory of future cases under current policy can help decision-makers assess whether mandates should be tightened or loosened [36, 34]. In practice, our model could be used to simulate possible outcomes by using the trained *β* network, but changing *M* to a time signal representing total lifting of restrictions, gradual reopening, or continuing heavy restriction. Such a model may need to account in some way for the costs of each policy.

Mixed machine learning models need not supplant traditional models entirely, but they can be a valuable auxiliary. As our model shows, they need not be overly complex or computationally expensive. They can interpret large amounts of data, generalize well to a variety of different regions and, given appropriate learning biases, can be relied upon to make feasible predictions.

Epidemic models are often under-determined by data [63]. UDEs allow a new approach to this problem. Since neural networks are universal approximators [64], they can represent the full range of possible functions that could fit the available data. By training multiple iterations of a UDE model and analyzing their trajectories, we can see a range of feasible outcomes for the system with just one single model. For example, two UDE predictions can fit the data and biological constraints equally well, yet one may predict a massive second wave, while the other predicts a rapid return to normalcy. A third may produce several smaller waves with corresponding mobility changes. That said, it is important to assign sufficient weight to the learning biases to avoid discouraging such a range of behaviours in favour of a single, overfitted solution.

The ability of UDEs to examine a range of data-fitting functions could be further enhanced with sparse regression methods [51, 65, 66]. By applying sparse regression to our trained *β* model’s output, one could derive a multitude of symbolic equations that could be used to mathematically model the system.

The results also support our hypothesis that learning biases are effective at accelerating training and assuring socially and biologically plausible solutions while achieving superior training performance. While some attributes can be learned passably well by the unbiased model given sufficient training time, the biased model still achieves better losses on these attributes by at least two orders of magnitude. Good performance on training data should not be taken too seriously since it may be a sign of overtraining. However, this does not appear to be the case in our model. The vast majority of the average training loss comes from a few highly divergent solutions. The improved performance by the biased model indicates reduced proclivity for such solutions.

The fact that a monotonic *β* is learned comparably well by both models indicates that both of them are instrumentally useful for the model to learn when satisfying the loss objective. This gives a good sanity check that these features are present in the real system and assuming them in the model is reasonable. The upper and lower bounds on mobility, however, are not typically inferred by the model without explicit instruction. This is not unexpected since the observed data never nears the bounds. By fitting the data well, the model never needs to learn what happens at those bounds. However, including these boundaries as learning biases gives greater assurance that the model will not produce divergent or unstable solutions if, for example, it were used to predict what would happen in a scenario where those bounds were neared. Of course, it is preferable to ensure stability mathematically using structural biases, but this may not always be feasible. The other objective which the unbiased model tends not to learn is in the tendency for *M* to return to the baseline value of 0. This may be because, as people become accustomed to life with the virus, the ”baseline,” i.e. the average societal preference in the absence of disease, shifts downward.

It is interesting that the learning biases help generate greater variability in the out-of-sample time series predictions. This is likely because in the absence of any other objective, the model consistently converges to a single global optimum for data fitting. Since the model’s extended prediction tends to remain within a small region of state space (*M* remaining negative, *I* relatively small), the greater potential variance is never realized. The model with learning biases, meanwhile, is relatively less concerned with fitting the data and hence has more freedom to explore the parameter space. The fact that the constraint losses are evaluated according to randomly generated sample points also confers greater variability to the results of the biased model.

The biased model also has greater variability in the upper quartile of its *β* response, but reduced variability in the lower quartile. This makes sense – the biased model has learned that for any *M* greater than those it has seen, the value of *β* must also be greater (and vice-versa for *M* less than what it has seen). The unbiased model, having no such information, cannot make an informed prediction, and so is equally likely to predict continued increase or an unrealistic decrease.

These variability trade-offs favour the biased model. Greater variability in time series prediction is valuable because (assuming the predictions are biologically feasible) it shows a greater variety of possible outcomes and assigns a degree of confidence associated to those outcomes. The reduced variability in predicting the transmissibility is also desirable because it derives from better understanding of the system.

We used a heavily simplified model of COVID-19. It is not intended to capture all details of the pandemic, nor is it meant to recommend specific health policies. We assumed the acquired immunity is permanent, which it may well not be [67]. We do not account for vaccines, which came into play around the end of 2020 [68]. Thus, the long-term predictions (i.e. beyond 300 days or so) should be taken only as evidence that the model does not produce wildly implausible behaviour, rather than a serious attempt to forecast cases too far in the future. The emergence of new variants, first reported in September 2020 [69] at the end of the second wave in many populations, mean that predictions for the tail end of 2020 are beyond the model’s intended scope.

Even in the short term, the model is not intended to predict cases or to precisely estimate the virus’s basic reproduction number. It is limited by our ability to consistently measure recovery rates and estimate under-reporting ratios, which almost certainly vary between regions and over time within regions. For simplicity, we also left out asymptomatic transmission and seasonal changes in infectiousness, both of which hamper the model’s short-term predictive ability compared to a more complex model [63]. None of these limitations changed our conclusions, since our goal was to show that UDEs and PIML can fit available data while making qualitatively correct our-of-sample predictions.

Future work could improve our the model by incorporating some of the aforementioned details of the pandemic. This could give insight into other behaviour-disease interactions like vaccine usage [70] or allow an examination of how these dynamics changed over the course of the pandemic. In section 4.1-4.2.1 we also provide some methodological changes that could further develop the UDE/PIML themes, particularly regarding how to use learning biases effectively.

Probably the biggest opportunity for future work is to apply this type of data-driven differential equation model to other systems. Other infectious diseases, particularly those for which vaccines are available, are also coupled behavior-disease systems [70, 66] and so could be amenable to this type of model. Beyond epidemic modelling, climate systems are also known to have important behavioural components [71, 72]. Ultimately, one of the greatest advantages of UDEs is that, as per their name, they can theoretically be applied to any dynamical system [51]. It is only a matter of testing them to see if they provide valuable insight which.

Availability of social and epidemiological data for endemics and future pandemics will likely continue to increase. At the same time, socio-economic factors will continue being complex, and regional and temporal variability will persist. Universal differential equations, when supplied with appropriate learning biases, could be a valuable tool for modelling such systems. Alongside traditional models, they could be used to quickly gain perspective on the state of outbreaks across the world without having to develop specialized models for each region.

## Materials and Methods

Our model is a universal delay differential equation (UDDE) based on the standard SIR model:

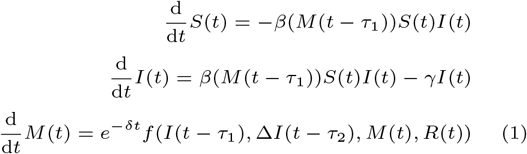

where *S, I* and *R* represent the susceptible, infected and recovered proportions of the population respectively (*R* can be recovered as 1 *− S − I*), and *M* represents the percent difference in mobility compared to the baseline. *β*(*M*) represents the transmission rate as it depends on mobility, and *f* (*I*, Δ*I, M, R*) represents the dynamics governing social/behavioural (mobility) response to the infectious disease [35]. Both *β* and *f* were learned by the algorithm. Δ*I*(*t*) represents the change in *I* between the current time *t* and a previous time *t − τ*_2_. The *e*^*−δt*^ factor accounts for several factors that reduce the population response to the virus over time, including: pandemic fatigue, the development of medical interventions that make the infection less fatal (such as ‘proning’), substituting less disruptive interventions (such as masking) for mobility reductions, and (for longer-term predictions than we study in this model) the evolution to milder virulence over time. Section 4 Figure 4 shows a schematic of the model.

**Fig. 4.**
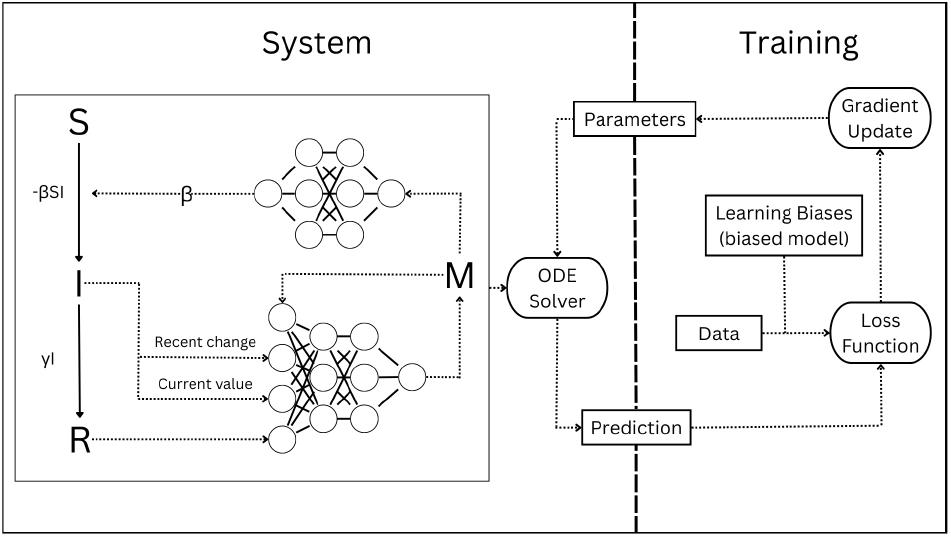
Schematic of our model showing the relevant differential equations, neural networks, and training procedure. The learning biases are present only in the biased model. Otherwise, the biased and unbiased models have the same structure.

The model parameters are *γ*, the per-capita recovery rate, *τ*_*M*_, the delay between a change in *M* and the corresponding change in prevalence, and *τ*_*R*_, the reverse delay: the time between a change in prevalence and corresponding behavioural response [35]. The values we used are *γ* = 0.25day^*−*1^ [73], *τ*_*M*_ = 10days, and *τ*_*I*_ = 14days [74, 62].

This model inherits several structural biases from the standard SIR model template. First, *S* = 0 and *I* = 0 are both invariant, preventing any unbiological negative values for these variables. Second, it retains the conservation relation *S* + *I* + *R* = 1. Thus, regardless of the functions fit by the neural network, *S*(*t*) and *I*(*t*) are guaranteed to be plausible.

### Neural Networks

The influence of mobility (i.e. contact rate) on the transmission rate is represented by neural networks that are used to represent *β*(*M*) and *f* (*I*, Δ*I, M, R*). These networks each have linear output layers with one neuron and 2 hidden layers with 3 neurons per hidden layer and Gaussian Error Linear Unit (GELU) activation functions. This gives the *β* network 22 parameters and the *f* network 31, for a total of 54 once *δ* is included. Different network structure (size, activation function) could have improved performance. The hyperparameter space is probably too large to globally optimize the model, but there may be improvements we missed in our hyperparameter tuning.

### Training Methodology

The baseline (unbiased) model, which received no social or biological feedback, was trained only to fit the data (details in section 4.3). The model’s prediction is generated by solving the delay differential equation system to get its prediction for each state at each time step. This prediction is then fed into a scaled mean-squared error loss function:

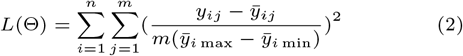

Here, *n* is the dimension of the system, *m* is the number of data points, *y*_*ij*_ is the true value of the *i*th variable’s *j*th data point, and 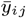 is the prediction for *i*th variable’s *j*th data point. *k* is the size of the parameter vector Θ, and Θ_*l*_ is the *l*th entry in Θ. Scaling the loss function in this way helps ensure all variables are given equal importance despite having different ranges [75].

Both biased and unbiased models for all regions were trained on the first 160 days, giving *m* = 22 data points after weekly averaging (see 4.3). This time period fully encompasses the first wave for all populations studied, but does not include the beginning of the second wave.

### Learning Biases

The socially and biologically informed model was trained to minimize the same accuracy loss objective as well as 8 other objectives, each encoding a social or biological assumption. These BIML loss functions are deliberately constructed so as to give 0 loss to any functions which satisfy the relevant assumptions. This allows the model complete freedom to explore the range of biologically feasible functions.

At each training iteration, the model generates 100 random points in the region 0 *≤ I ≤* 1, *I −* 1 *≤* Δ*I ≤ I, −*100 *≤ M ≤* 100. The BIML loss functions are evaluated at each of these points. The total loss at each iteration is then a weighted sum of the BIML losses, plus the accuracy loss. We tried dynamically updating the weights for each loss function as in [25], but this did not significantly improve results. The BIML assumptions and corresponding loss functions are displayed in Section 4.2.1 Table 3.

**Table 3.**
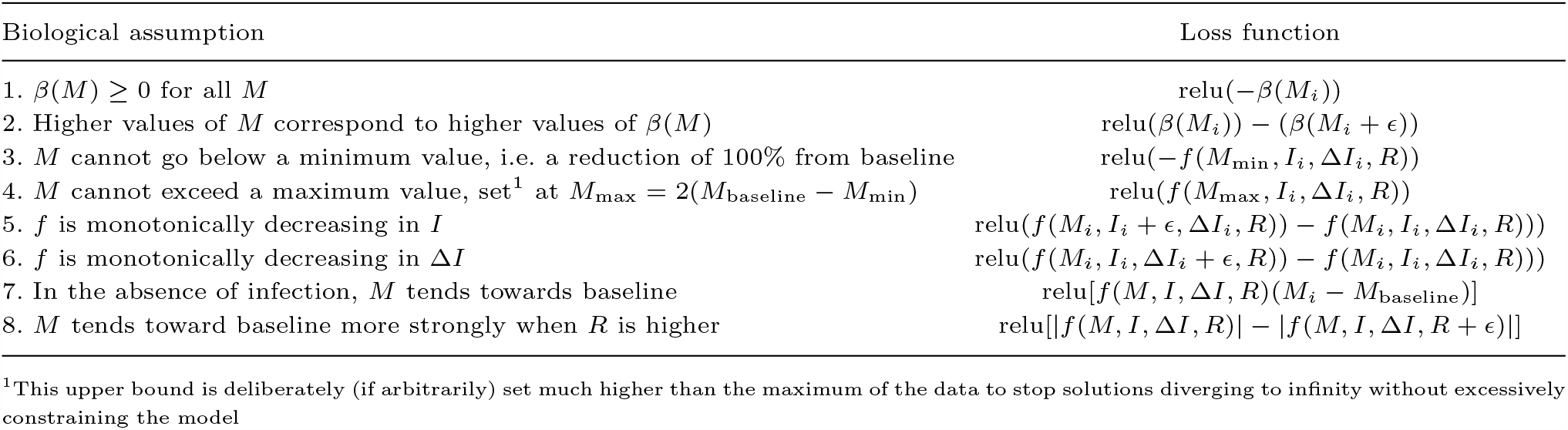
BINN loss functions

We only tested a few values for the learning bias weight. The optimal value for achieving tolerable performance on training data while assuring qualitatively realistic long-term predictions may be higher or lower. It may instead involve weights that differ between loss functions or which evolve over the training process.

Model parameters were randomly initialized. Parameter sets that gave initial errors of more than 10^4^ were re-initialized. We used the Adam optimizer to optimize the model parameters. For the basic model, we simply use the gradient of the accuracy loss with respect to the parameters. For the biologically-informed model, we use the same weighted sum for the total gradient as for the total loss. More details on the software implementation of this algorithm can be found in the appendix.

We found that training the model on the entire training set at once caused it to become stuck in a local optimum where *I* never increased. Thus, we trained the models in stages to achieve a better fit more quickly. The model trained on the first quarter of the data in the first stage, the first half in the second, and the entire training set in the third.

Repeating our model with more computing time and power could be informative. Although we were able run the model with enough iterations to ensure all models converged to a good degree, some certainly converged better than others. The mobility data was a particular challenge, with fairly sharp downturns and upturns occasionally not always fully captured. This could be assisted by using collocation-based training to speed up the process [76].

## Data

### Case Data

Daily case notification data was taken from the Johns Hopkins CSSE dataset [77]. We derived daily susceptible and infected proportions using the following system:

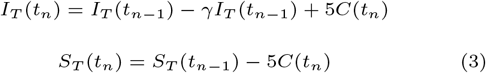

where *S*_*T*_ (*t*_*n*_) and *I*_*T*_ (*t*_*n*_) are the total number of susceptible and infected individuals (not proportions) on day *t*_*n*_ and *C*(*t*_*n*_) are the number of new cases on day *t*_*n*_. The parameter *γ* is the same used in the model as discussed above. Finally, we divided *I* and *S* by the total population to get a proportion at each time step to ensure different regions are comparable.

### Mobility Data

Daily mobility data (*M*) comes from the Google Community Mobility Report [78]. We mean-normalized this data to give it a comparable range to S and I. We chose the Retail and Recreation category of mobility data as it corresponded most to what we were trying to measure: voluntary activities in indoor settings that place people at risk of becoming infected. It is also strongly correlated with infection, so it is reasonable to expect it can be a good predictor of cases. Repeating the simulations with workplace mobility would be a good test of the model’s validity. Other mobility measures, such as parks, would be difficult to use due to their weaker correlation with infections [17].

Once we had data for each day, we sub-sampled it by taking a weekly moving average to reduce irregularities from weekends, holidays, and days where data was not available. We set the initial condition as the first data point for which *I* ≠ 0.

The regions analyzed were chosen to represent a variety of relatively populous locations across Western Europe, the US, and Canada. We chose to focus particularly on regions where populations are decently concentrated and which had notable second waves.

## Data Availability

All data produced in the present study are available upon reasonable request to the authors

https://github.com/bkuwahara/predicting-2ndwaves-udes

## Acknowledgments

The authors thank the anonymous reviewers for their valuable suggestions.

## Supplementary Material

Supplementary material is available at PNAS Nexus online.

## Author contributions statement

C.T.B. conceived the project, provided guidance and direction, conducted and shared preliminary research, and reviewed the manuscript. B.K. designed the specific details of the model, coded and ran the simulations, and analyzed the results. Both authors contributed to writing the manuscript.

## Data availability

The code used to run and analyze the model is available at https://github.com/bkuwahara/predicting-2ndwaves-udes. All empirical data are publicly available (see References).

