## Supplementary Appendix for "Predicting COVID-19 pandemic waves with biologically and behaviorally informed universal differential equations"

**Abstract**

In the early stages of the COVID-19 pandemic, it became clear that pandemic waves and population responses were locked in a mutual feedback loop. The initial lull following strict interventions in the first wave often led to a second wave, as restrictions were relaxed. We test the ability of new hybrid machine learning techniques, namely universal differential equations (UDEs) with learning biases, to make predictions in a such a dynamic behavior-disease setting. We develop a UDE model for COVID-19 and test it both with and without learning biases describing simple assumptions about disease transmission and population response. Our results show that UDEs, particularly when supplied with learning biases, are capable of learning coupled behavior-disease dynamics and predicting second waves in a variety of populations. The model predicts a second wave of infections 55% of the time across all populations, having been trained only on the first wave. The predicted second wave is larger than the first. Without learning biases, model predictions are hampered: the unbiased model predicts a second wave only 25% of the time, typically smaller than the first. The biased model consistently predicts the expected increase in the transmission rate with rising mobility, whereas the unbiased model predicts a decrease in mobility as often as a continued increase. The biased model also achieves better accuracy on its training data thanks to fewer and less severely divergent trajectories. These results indicate that biologically informed machine learning can generate qualitatively correct mid to long-term predictions of COVID-19 pandemic waves.

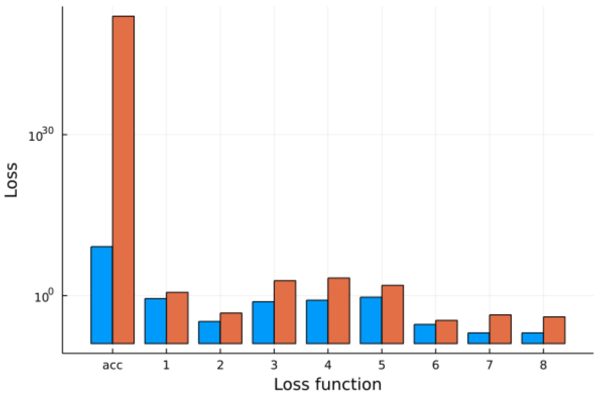


Figure 1: Trained model loss for the biased model (blue) and unbiased model (orange). Loss functions are indexed as in table 3 of the main paper

Table 1: Predicted $R_0$ and required mobility reduction (unbiased model)

| Region | Predicted $R_0$ | $M_{crit}$ (95% credible interval) | Convergence [a] |
|---|---|---|---|
| Austria | 2.1 (±0.38) | −0.32 (±0.35)[b] | 0.94 |
| Belgium | 2.5 (±1.3) | −0.32 (±0.62)[b] | 0.8 |
| Germany | 2.5 (±1.71) | −0.37 (±0.27) | 0.92 |
| Italy | 2.1 (±1.3) | −0.41 (±0.44)[b] | 0.89 |
| Netherlands | 2.0 (±0.36) | −0.43 (±0.15) | 0.97 |
| UK | 1.9 (±0.53) | −0.37 (±1.32)[b] | 0.70 |
| California | 1.5 (±2.1) | −0.24 (±1.73)[b] | 0.41 |
| New York | 2.3 (±0.11) | −0.58 (±0.31) | 0.97 |
| Pennsylvania | 2.4 (±0.26) | −0.43 (±0.59)[b] | 0.63 |
| Texas | 1.8 (±0.59) | −0.55 (±0.29) | 0.54 |
| British Columbia | 1.6 (±0.52) | −0.31 (±0.42) | 0.89 |
| Ontario | 2.5 (±1.3) | −0.47 (±10.90)[b] | 0.90 |
| Quebec | 2.4 (±1.1) | −0.32 (±0.46) | 0.92 |

[a] Not all neural networks showed a root. Interval calculated using only those that did.
[b] Interval exceeds physically realistic values

### 1 Biased vs unbiased comparisons: all regions

#### 1.1 Time series predictions

Section 1.1 figures 3 - 13 show the predicted time series of all model states for each region (except New York, which can be found in the Main Text, Section X Figure X) with learning biases (panels a-c) and without (panels d-f). Panels (a) and (b) show susceptible fraction, (b) and (d) show infected, and (c) and (f)

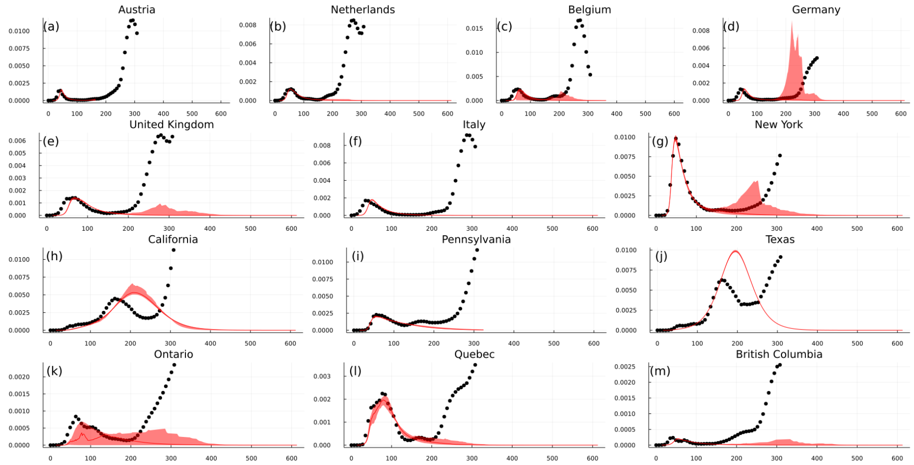


Figure 2: Infection prevalence time series predictions for all regions produced by the model with learning biases. Infection prevalence is the proportion of the total population that is infected at any given time. Green dots represent training data (first 22 weeks) and black dots show unseen data (a further 23 weeks). Predictions are generated using the median (solid line) and interquartile range (ribbon) of 100 independently-trained instances of the model per region.

show mobility. Green dots represent training data, while black dots represent unseen data. All predictions are generated using the median (solid line) and interquartile range (ribbon) of 100 independently-trained instances of the model.

#### 1.2 Transmissibility response

Section 1.2 figures 14-24 show predicted force of infection based on mobility level for all regions (except New York, which can be found in the Main Text, Section X Figure X) with learning biases (a) and without them (b). Dotted lines indicate values of mobility seen by the model during training. Solid line shows the median prediction of 100 model instances and ribbon shows interquartile range.

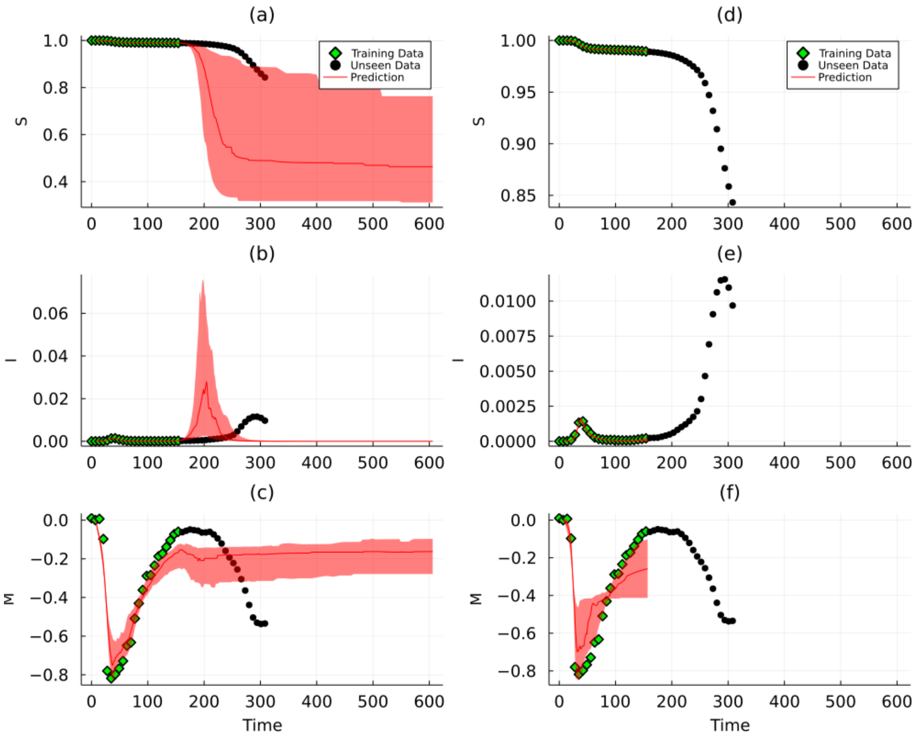


Figure 3: Time series comparison for Austria

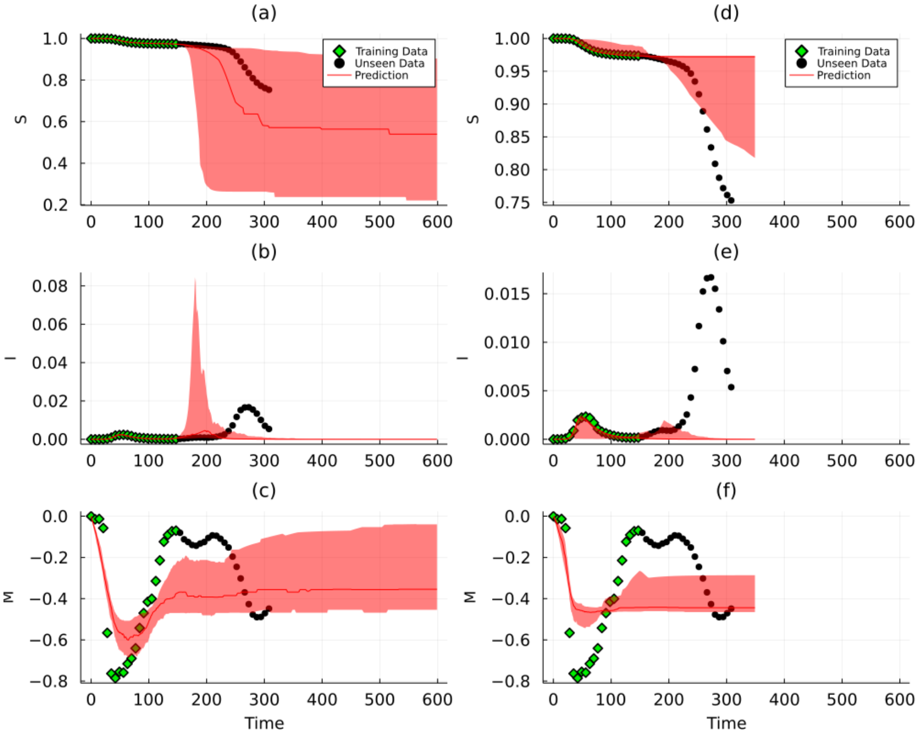


Figure 4: Time series comparison for Belgium

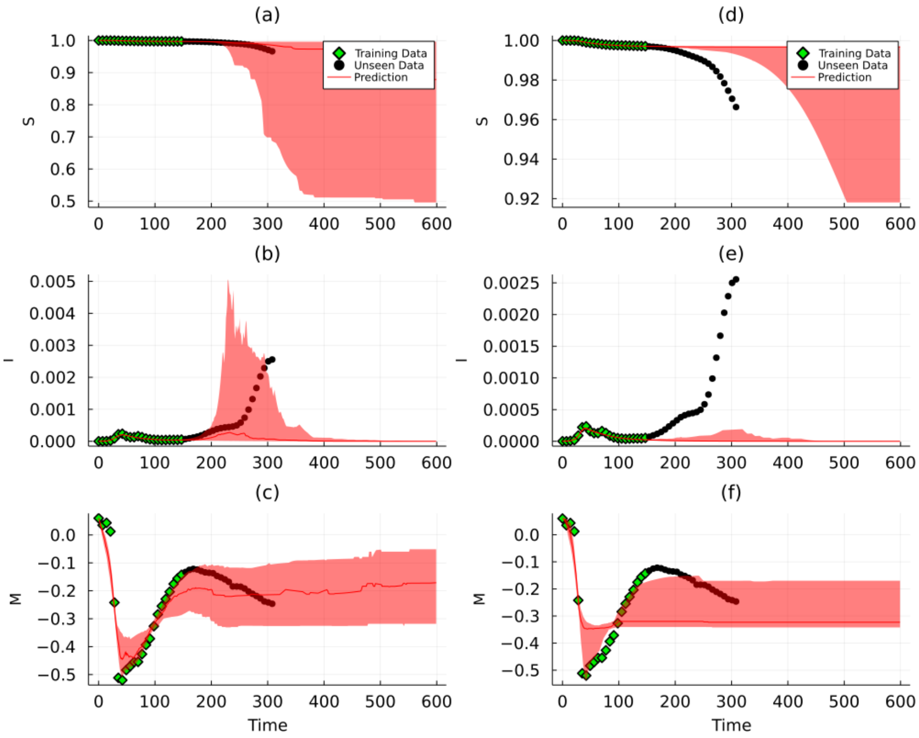


Figure 5: Time series comparison for Italy

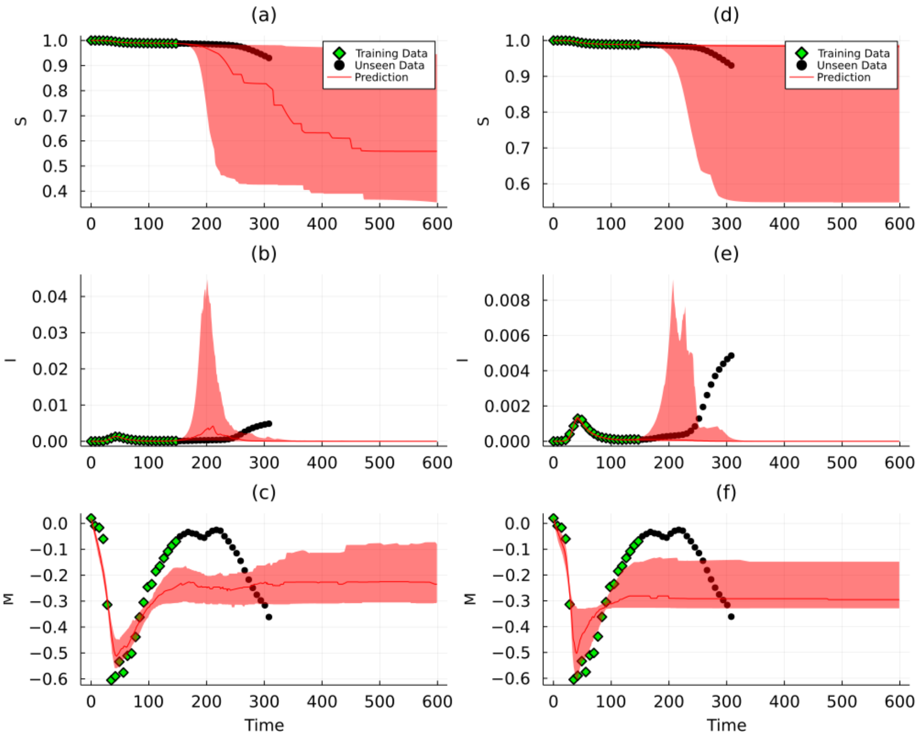


Figure 6: Time series comparison for Germany

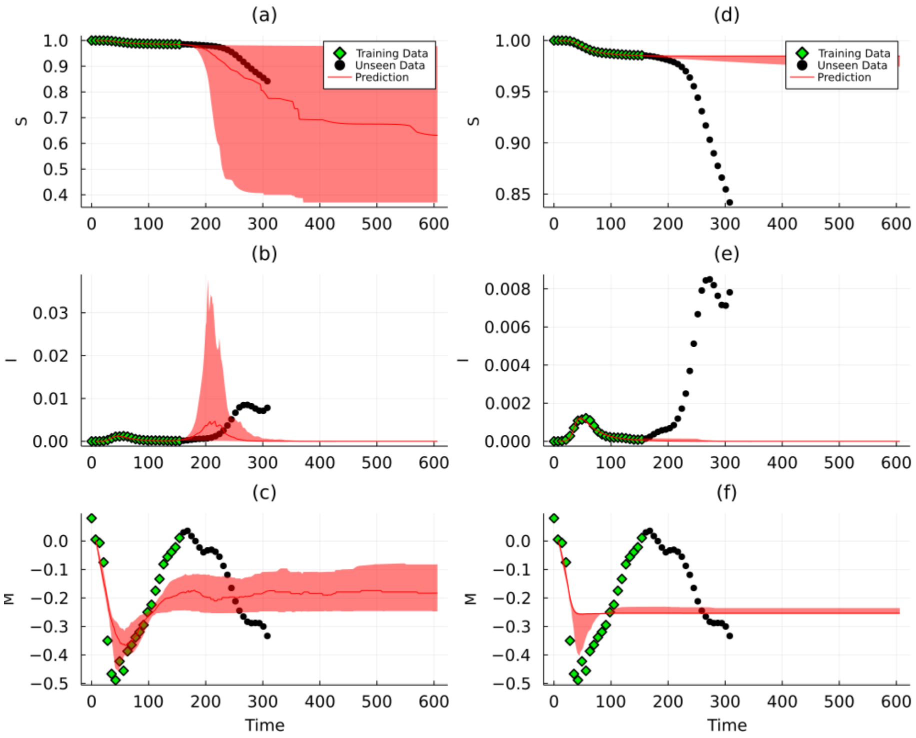


Figure 7: Time series comparison for Netherlands

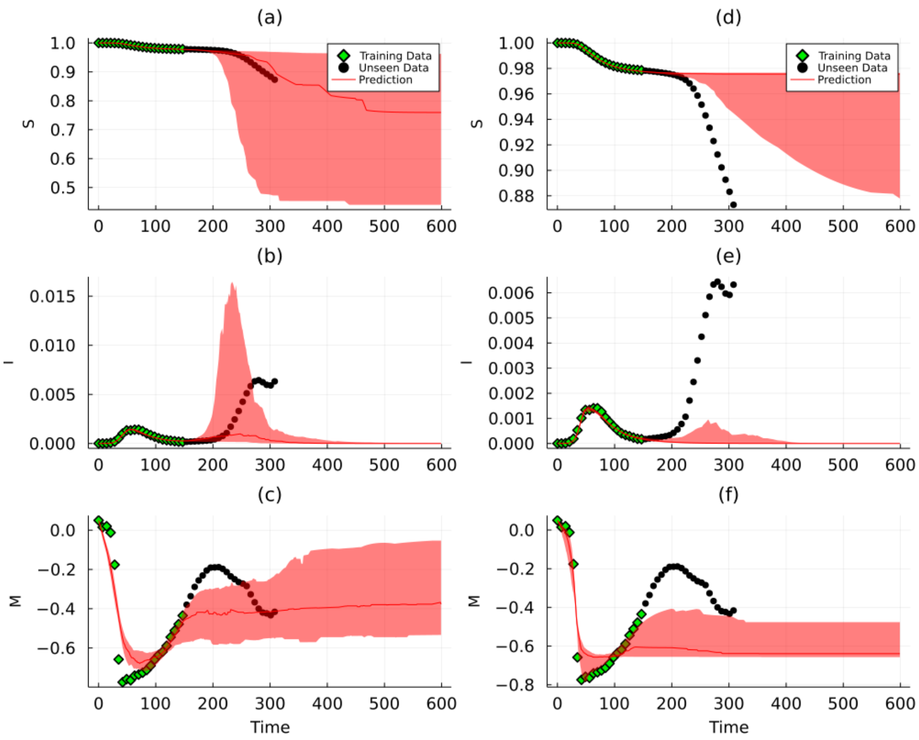


Figure 8: Time series comparison for the United Kingdom

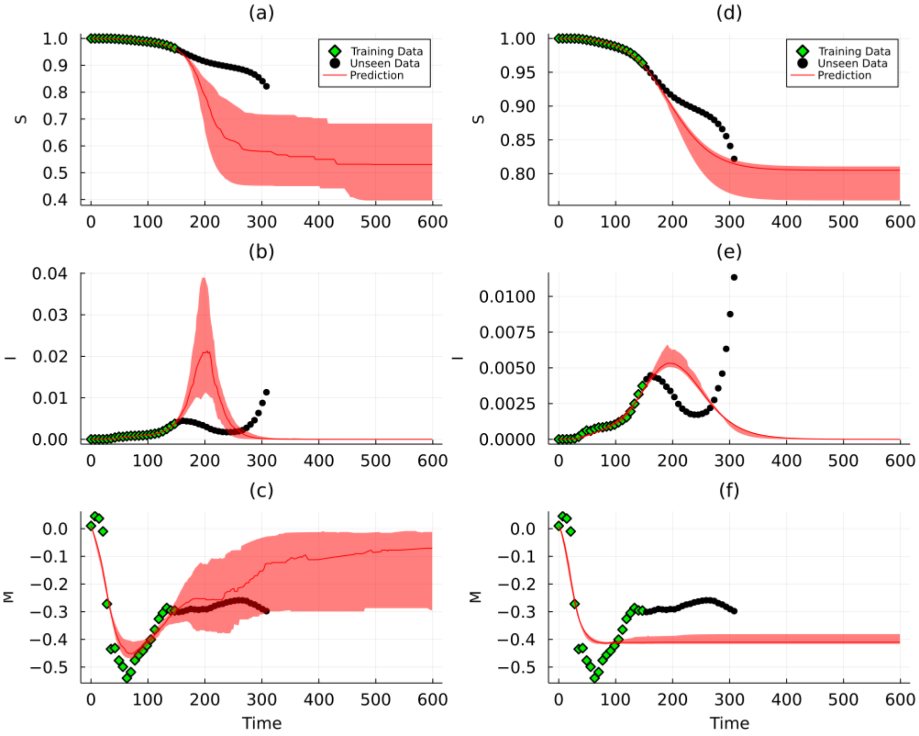


Figure 9: Time series comparison for California

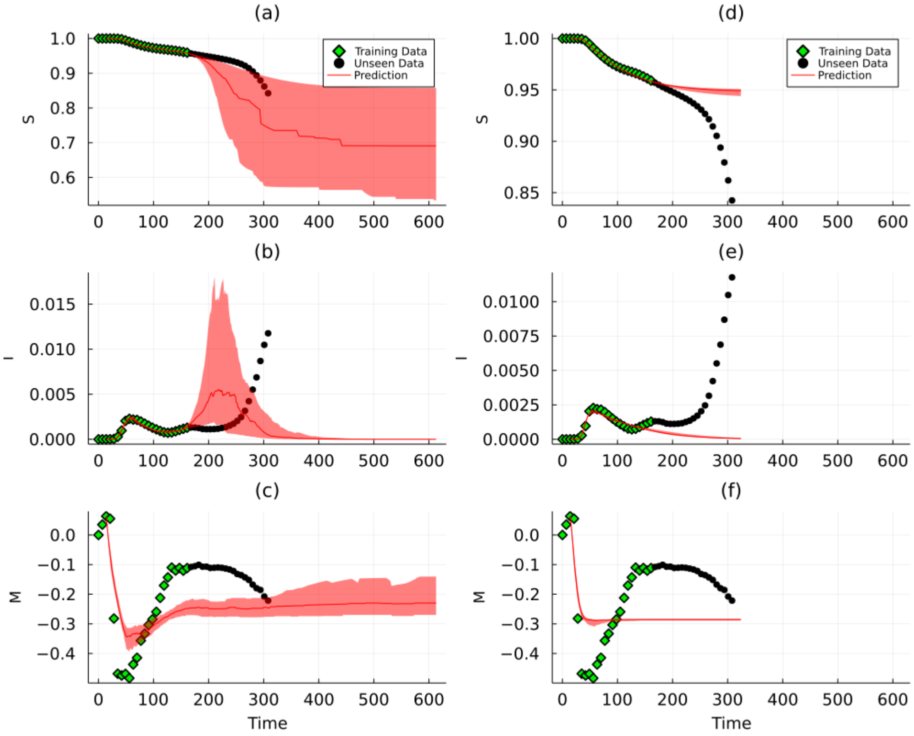


Figure 10: Time series comparison for Pennsylvania

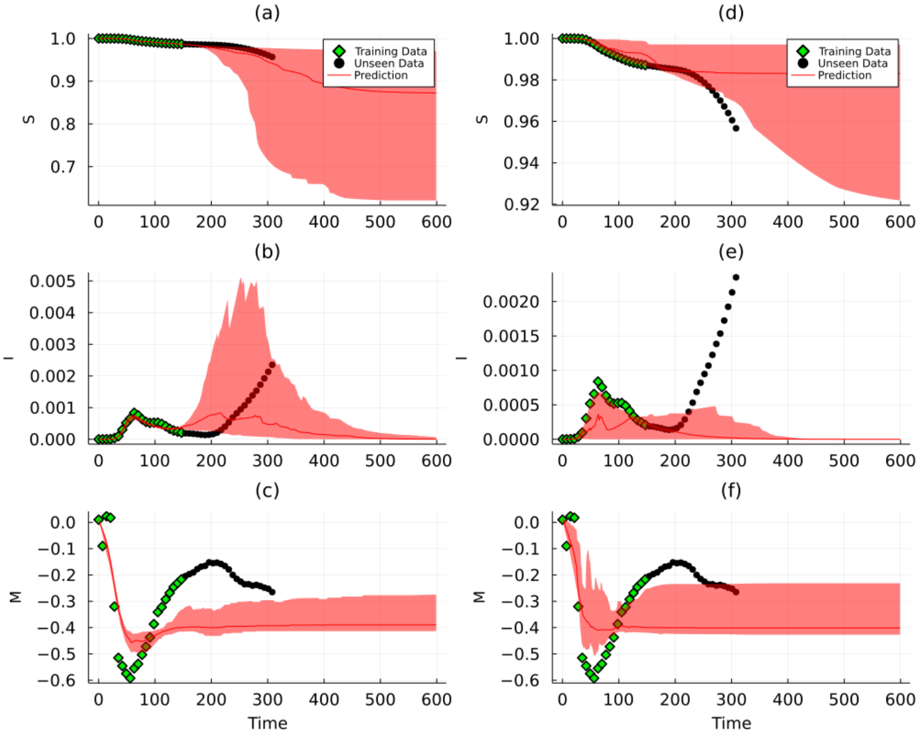


Figure 11: Time series comparison for Ontario

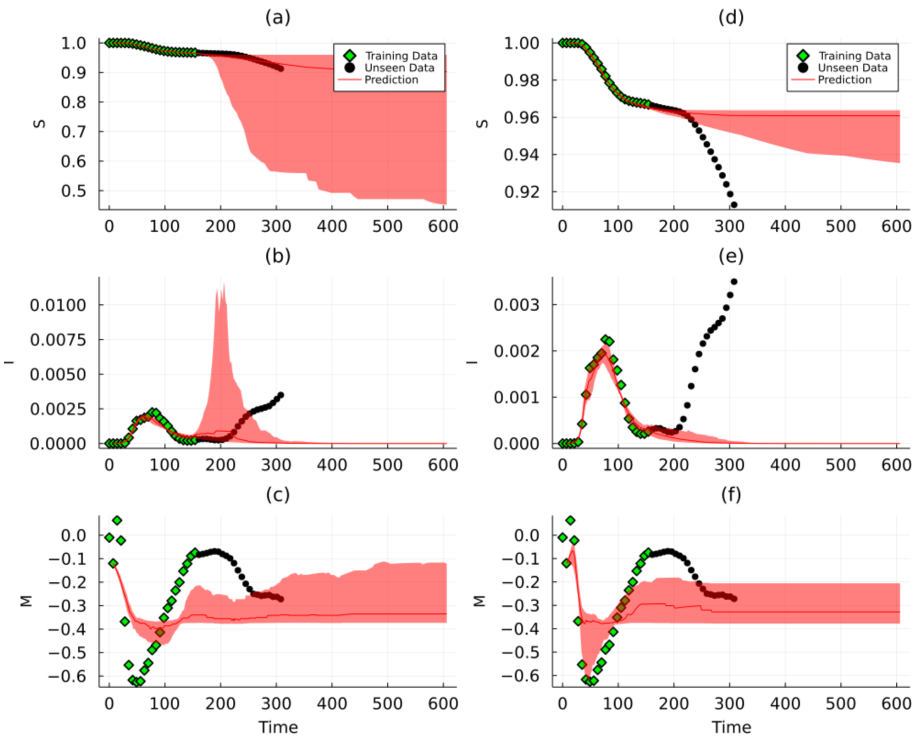


Figure 12: Time series comparison for Quebec

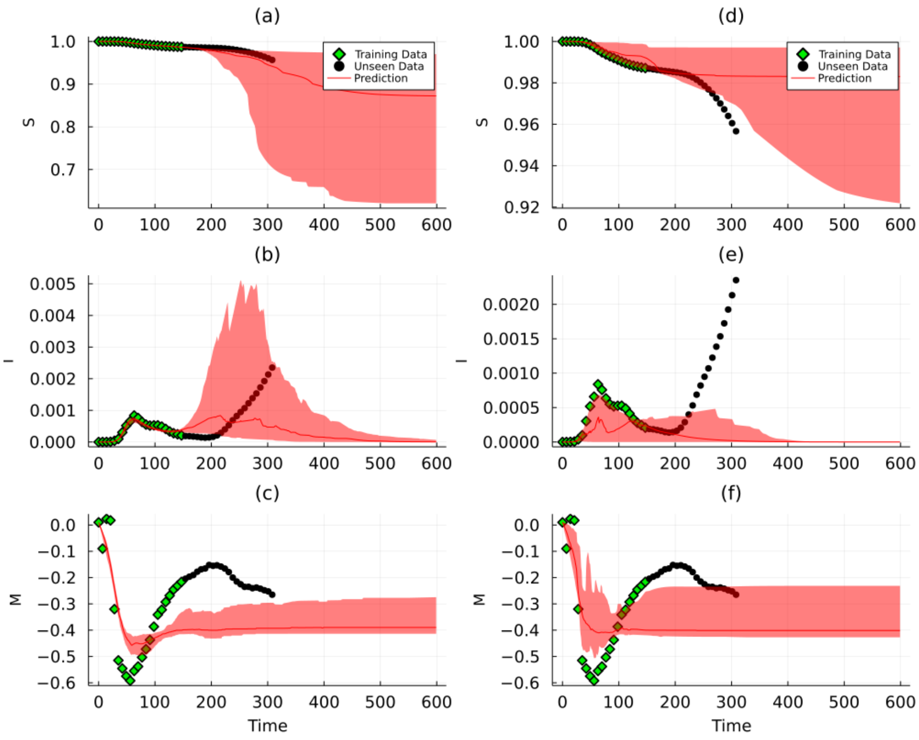


Figure 13: Time series comparison for British Columbia

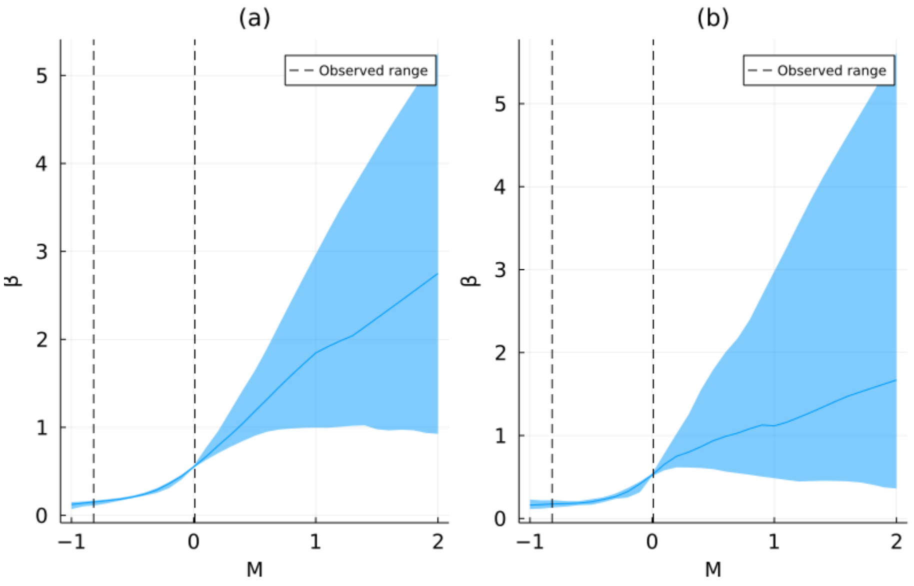


Figure 14: Transmissibility Response for Austria

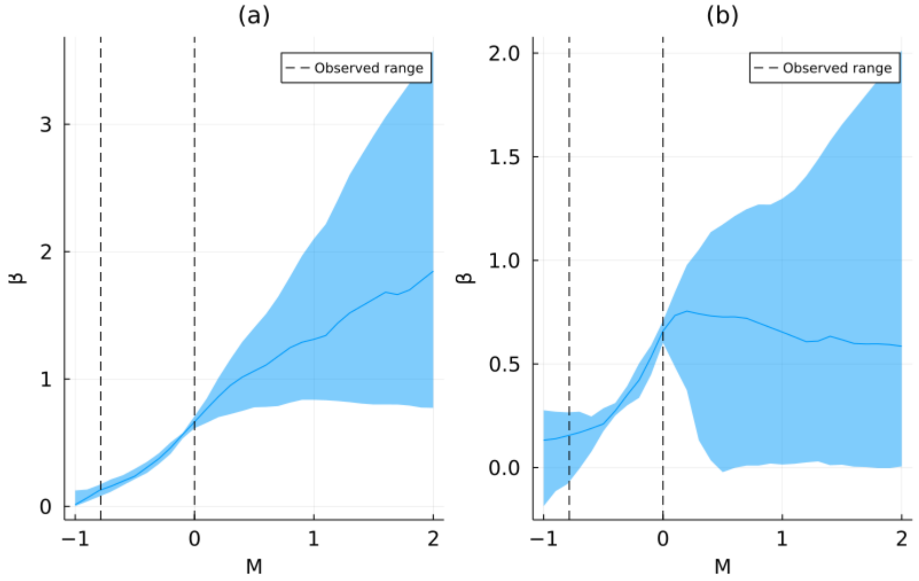


Figure 15: Transmissibility Response for Belgium

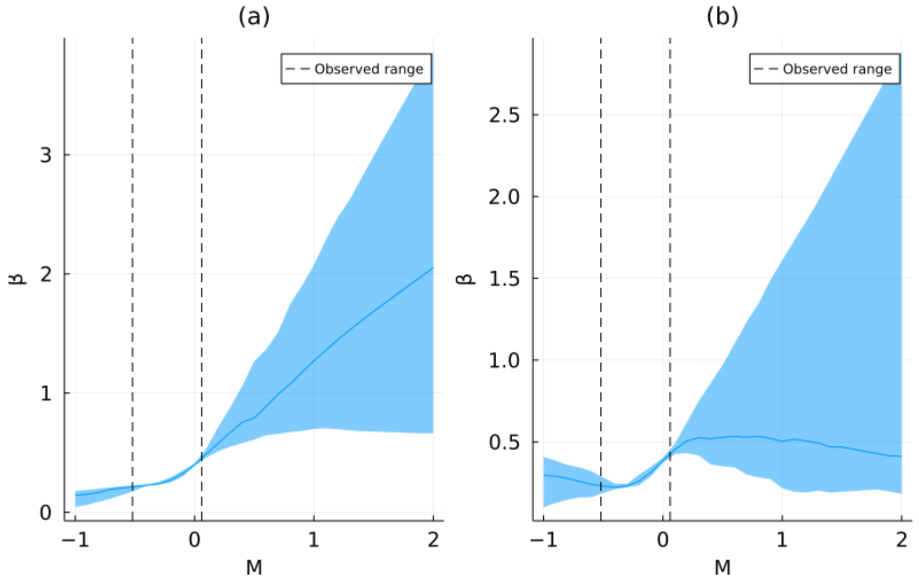


Figure 16: Transmissibility Response for Italy

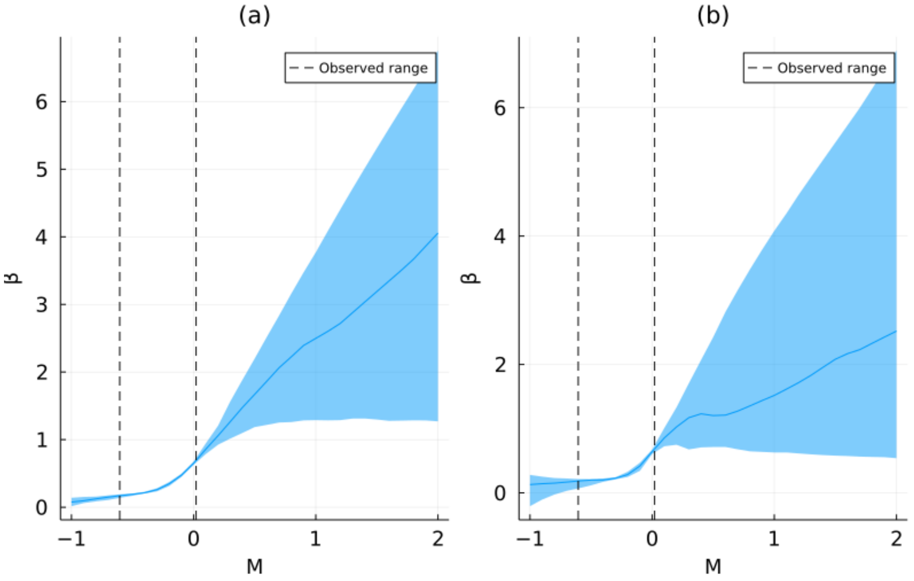


Figure 17: Transmissibility Response for Germany

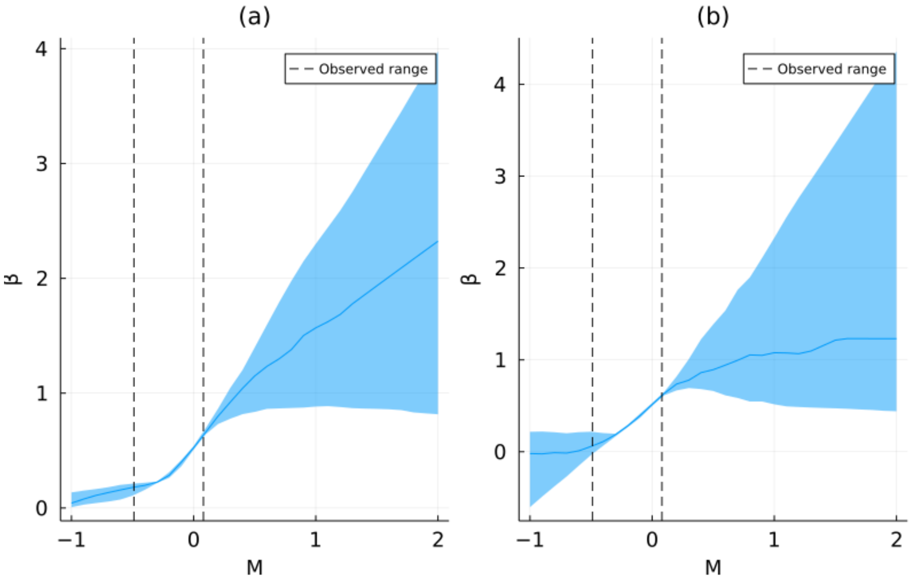


Figure 18: Transmissibility Response for Netherlands

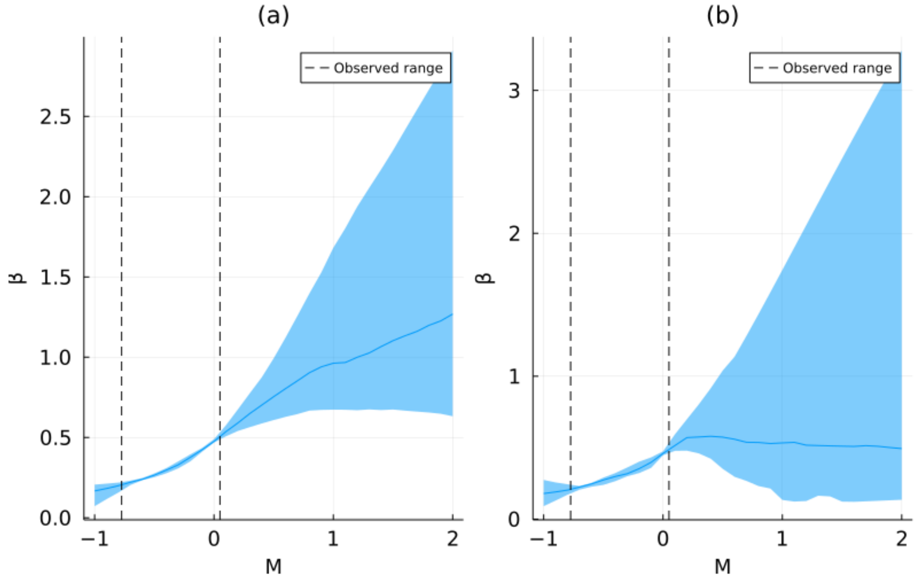


Figure 19: Transmissibility Response for the United Kingdom

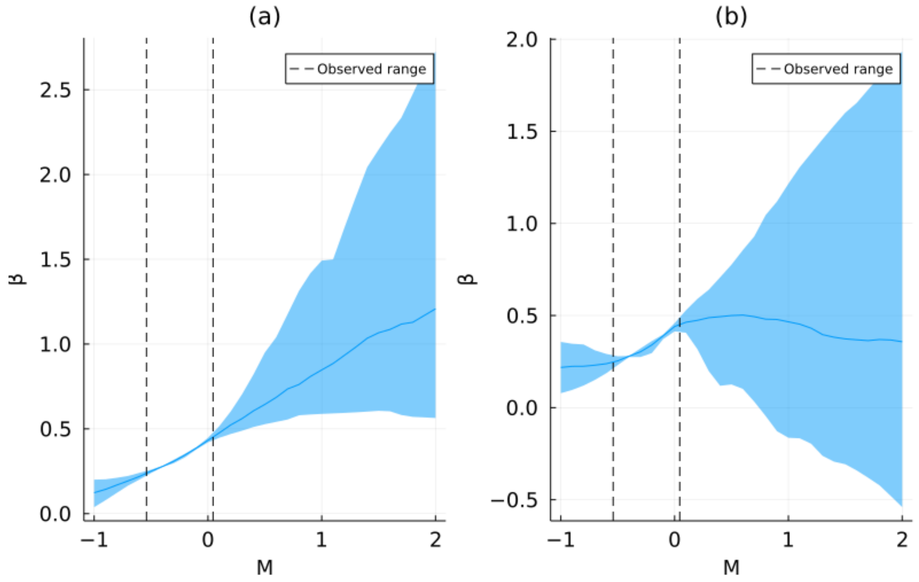


Figure 20: Transmissibility Response for California

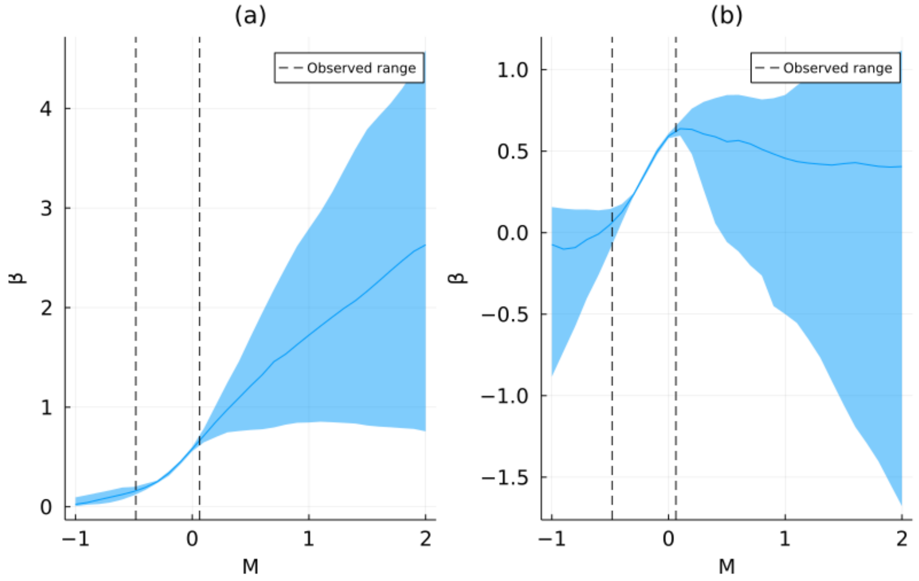


Figure 21: Transmissibility Response for Pennsylvania

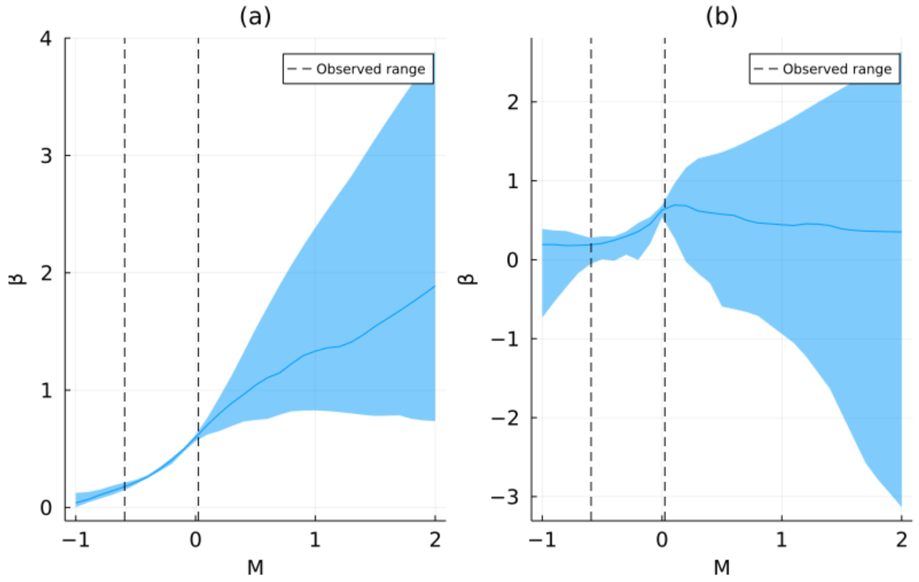


Figure 22: Transmissibility Response for Ontario

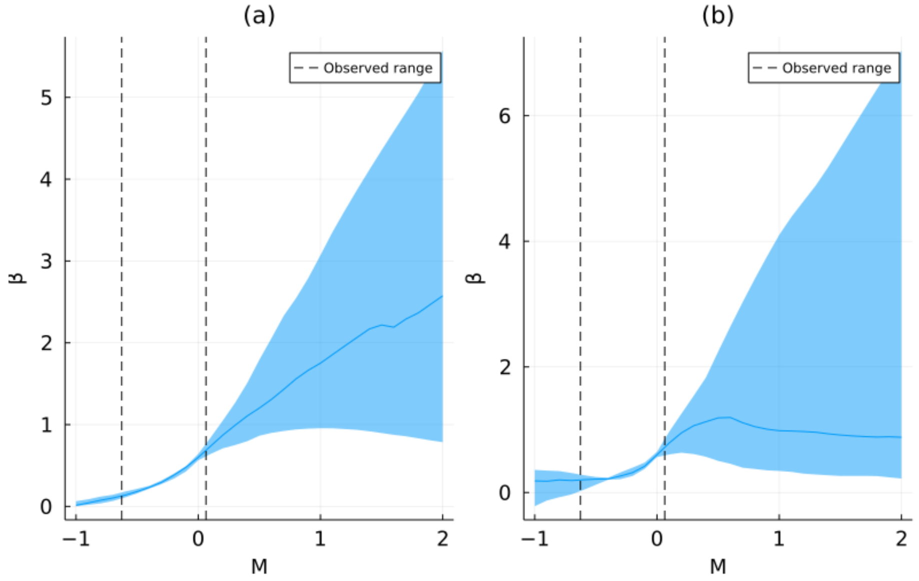


Figure 23: Transmissibility Response for Quebec

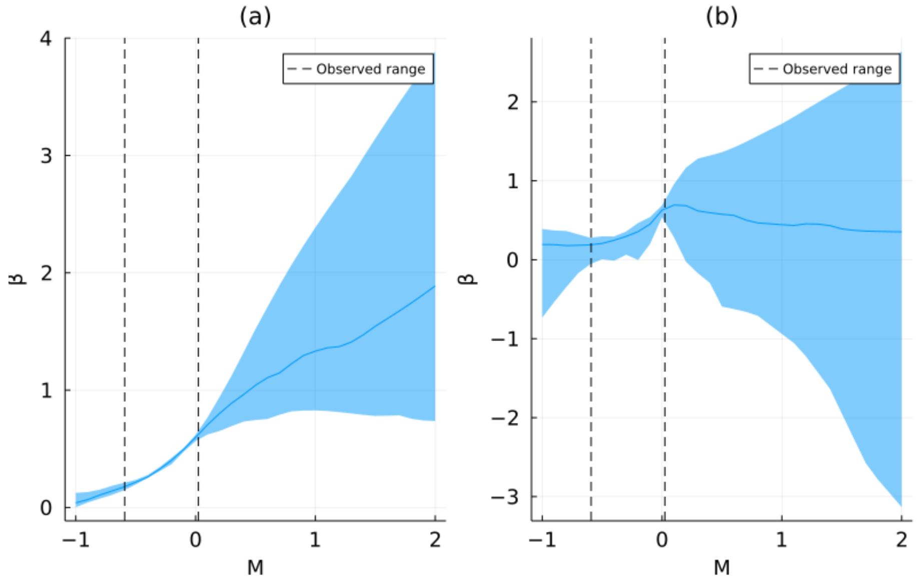


Figure 24: Transmissibility Response for British Columbia
